# Variation in Hospital Visiting Hour Policies in US Acute Care Facilities: An Exploratory Cross-Sectional Analysis

**DOI:** 10.64898/2026.04.27.26351861

**Authors:** Coby Y. Garcia, Cody Chou, Elizabeth Caso, James C. Hudspeth, Lao-Tzu Allan-Blitz

## Abstract

**Background:** Hospital visiting-hour policies vary widely across the United States, yet the structural factors shaping this variation remain poorly characterized.

**Objective:** This study investigates how hospital-level financial characteristics, payer mix, and rurality relate to the restrictiveness of inpatient visiting-hour policies, and assesses whether these relationships differ across states with diverse Medicaid expansion statuses.

**Design:** Cross-sectional observational analysis of hospital visitor policies in four states (Massachusetts, Wisconsin, Tennessee, and South Carolina) selected based on Medicaid expansion status, population size, and hospital density.

**Participants:** A total of 318 acute-care hospitals were included using publicly available data from the Centers for Medicare & Medicaid Services and the National Academy for State Health Policy.

**Main Measures:** The primary outcome was total daily visiting hours in general inpatient wards. Predictors included volume/capacity, patient mix, financial performance/efficiency, geography and organizational structure.

**Key Results:** Hospital-level characteristics including higher Medicaid payer mix, stronger financial margins, greater inpatient occupancy, and larger size were associated with shorter visiting hours in unadjusted analyses. Commercial payer mix and rurality predicted longer hours. Mean visiting duration was 14.1 hours/day (SD = 5.07; range 0–24), with Massachusetts having the shortest on average across states (10.5 hours/day) and Wisconsin the longest (16.3 hours/day). Medicaid payer mix was the only predictor associated with visiting-hour restrictiveness after multiple-testing correction. Each 10-percentage-point increase in Medicaid payer mix was associated with an approximately 11.3% decrease (p = 0.002) in visiting hours. Within-state variation exceeded the differences between-states.

**Conclusions:** Visitation hours vary considerably, with correlations around rurality of the community served, size of the hospital, and the number of patients on Medicaid. Medicaid payer mix emerged as the most consistent predictor of restrictiveness after adjustment. Hospitals can use these findings to evaluate visitation practices to balance patient-centered care with operational demands.

## INTRODUCTION

Hospital visiting hours dictate when patients can receive in-person support during inpatient care and influence patient experience and clinical outcomes, yet remain understudied. Evidence from the critical care context links more flexible visitation to better communication between families and care teams, lower patient anxiety, and shortened recovery times (1–5). Although much of this literature comes from intensive care settings, existent evidence in non-critical care populations suggests that family presence and fewer restrictions are associated with improved patient experience, reduced anxiety, and lower rates of delirium (6–8). U.S. hospitals vary widely in visitation policies, driven by differences in hospital type, staffing structures, safety protocols, and administrative norms, but the drivers of this variation are poorly understood (9). Some U.S. hospitals have removed any restrictions to visiting hours, with results reporting positive outcomes like fewer phone calls and greater family satisfaction (10,11). Hospital resources and organizational constraints may shape visitation policies. Facilities with limited staffing or tighter finances may restrict visitation to reduce perceived operational burden, whereas hospitals trying to improve patient experience may expand visiting hours (12). Concerns about increased disruption from visitors also impact approaches to visitor policy, potentially leading to more restrictive approaches, especially if there is a perception that some of the patient populations served by an institution have a higher risk of conflict (9).

Insurance reimbursement policy environments shape hospital payer mix, reimbursement levels, and fiscal stability, which can in turn impact staffing, the capacity to manage visitors, and administrative policies (13). As of October 2025, forty states and the District of Columbia have expanded Medicaid (14). Massachusetts, an early adopter of near-universal coverage (2006) and full expansion (2014), has one of the lowest uninsurance rates nationwide (<3% in 2022) (14–16). Wisconsin implemented a partial expansion in 2014, extending eligibility to 100% of the federal poverty level without the enhanced federal match, and reports a 5.90% uninsurance rate (14,17). Tennessee and South Carolina did not expand Medicaid and maintain higher uninsurance rates (9.6% and 8.2%, respectively) (14,18,19). Guided by Resource Dependence Theory, which posits that organizations adapt to external resource environments (20,21), we hypothesize that Medicaid expansion may create financial slack by reducing uncompensated care, enabling investment in patient-centered practices such as expanded visitation (22–25).

This study examines how hospital characteristics relate to the restrictiveness of visiting hours in U.S. hospitals. This study also explored whether those relationships differ across states that have or have not expanded Medicaid. Understanding the variation in visitor policies may inform efforts to optimize visitation policies and improve patient- and family-centered care.

## METHODS

### Research Design

This cross-sectional observational study examined associations between hospital visiting-hour policies and financial, structural, and payer-mix characteristics. States were selected to ensure: (1) heterogeneity in Medicaid expansion status and participation under the Affordable Care Act (ACA), (2) similar population size and (3) similar hospital density defined as inpatient hospital beds per 1,000 residents (Table 1). All acute-care hospitals with publicly available data were included from each state.

**Table 1.**
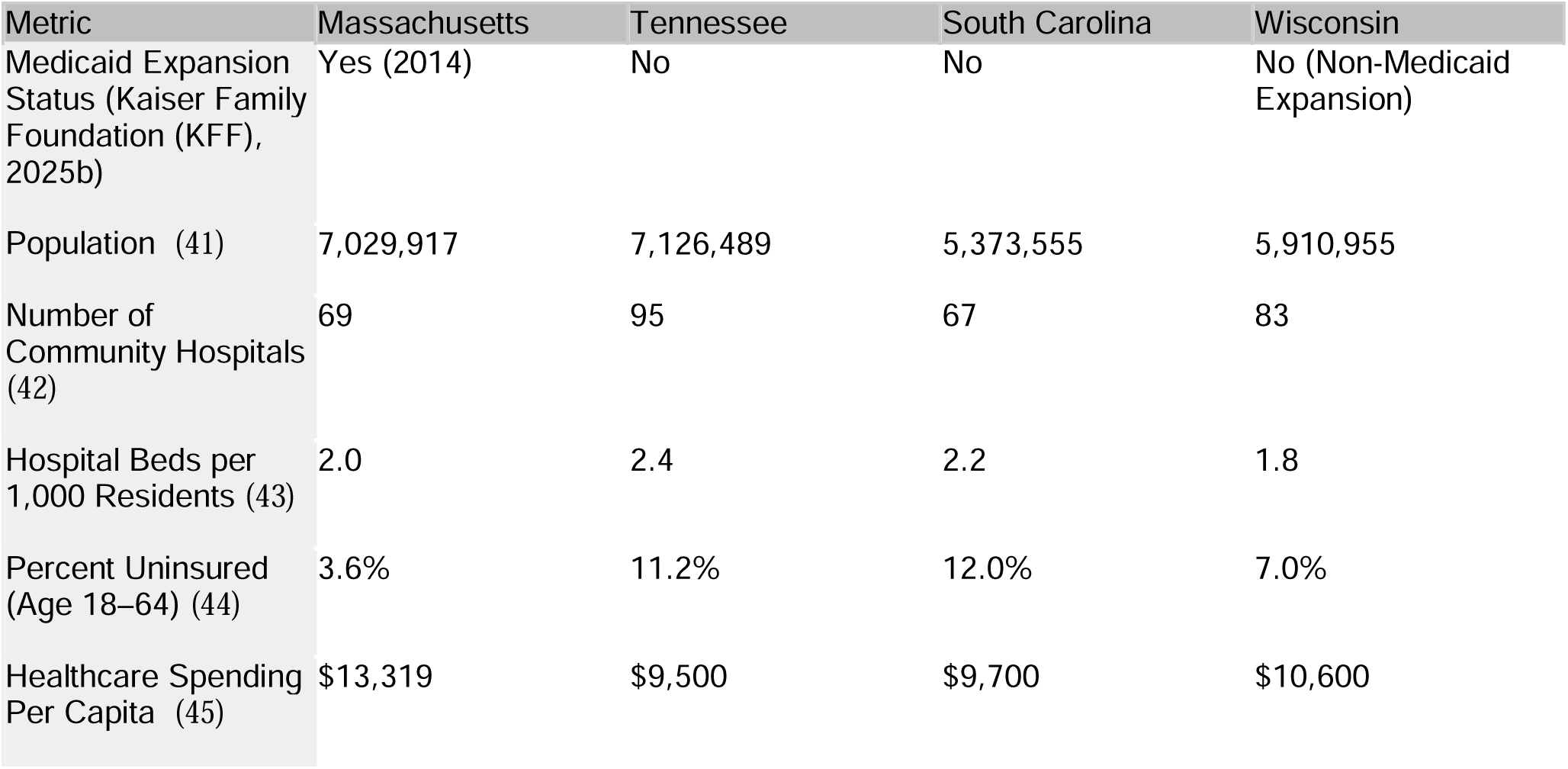
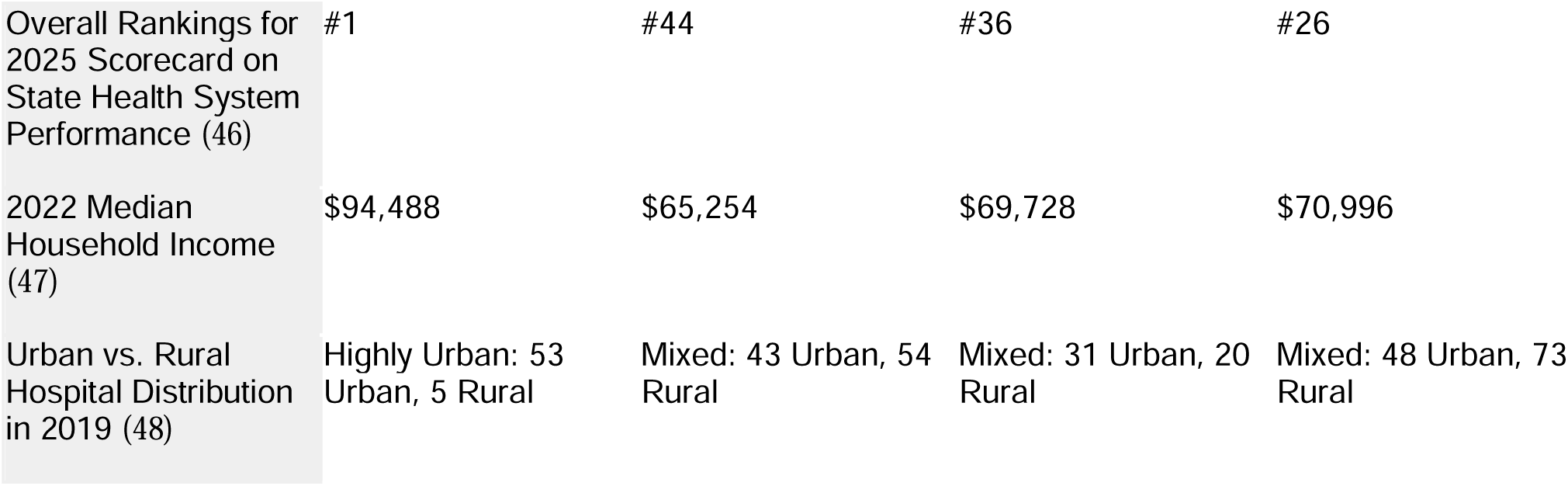
State-Level Demographic, Insurance, and Health-System Profiles for the Four Study States.

| Metric | Massachusetts | Tennessee | South Carolina | Wisconsin |
| --- | --- | --- | --- | --- |
| Medicaid Expansion Status (Kaiser Family Foundation (KFF), 2025b) | Yes (2014) | No | No | No (Non-Medicaid Expansion) |
| Population (41) | 7,029,917 | 7,126,489 | 5,373,555 | 5,910,955 |
| Number of Community Hospitals (42) | 69 | 95 | 67 | 83 |
| Hospital Beds per 1,000 Residents (43) | 2.0 | 2.4 | 2.2 | 1.8 |
| Percent Uninsured (Age 18–64) (44) | 3.6% | 11.2% | 12.0% | 7.0% |
| Healthcare Spending Per Capita (45) | \$13,319 | \$9,500 | \$9,700 | \$10,600 |
| Overall Rankings for 2025 Scorecard on State Health System Performance (46) | #1 | #44 | #36 | #26 |
| 2022 Median Household Income (47) | \$94,488 | \$65,254 | \$69,728 | \$70,996 |
| Urban vs. Rural Hospital Distribution in 2019 (48) | Highly Urban: 53 Urban, 5 Rural | Mixed: 43 Urban, 54 Rural | Mixed: 31 Urban, 20 Rural | Mixed: 48 Urban, 73 Rural |

### Study Population

The unit of analysis was the hospital. Data were compiled from five publicly available repositories: 1) Centers for Medicare & Medicaid Services Hospital General Information dataset, which provided structural, ownership, and quality metrics for all registered hospitals in the fiscal year of 2023; 2) National Academy for State Health Policy Hospital Cost Tool, which included financial indicators such as payer mix, operating margins, and cost-to-charge ratios for the fiscal year of 2023; 3) American Community Survey, which provided census-level estimates of poverty within hospital service areas for the fiscal year of 2023; 4) U.S. Department of Agriculture Rural-Urban Commuting Area Codes by ZIP Code, derived from the 2020 decennial U.S. Census which classified areas based on population density, urbanization, and daily commuting patterns from the 2020 census; and 5) Agency for Healthcare Research and Quality (AHRQ) Compendium of U.S. Health Systems (CHSP), which provided system membership and teaching status.

### Measures

The primary outcome was the total daily visitation hours in general inpatient wards, as both continuous and categories: highly restrictive (≤6 hours), moderately restrictive (>6–12), generous (>12–18), and open (>18–24) (26).

In univariate analyses, we evaluated a broad set of hospital-level predictors, including financial characteristics (Medicaid payer mix; combined government payer mix [Medicaid, CHIP, and other low-income programs]; commercial payer mix; Medicaid, operating, and net profit margins; operating costs; total operating expenses; cost-to-charge ratio; and net charity care cost as a percentage of revenue), capacity and utilization (bed size, inpatient occupancy, adjusted discharges), labor costs (direct patient-care labor cost per full-time equivalent and per adjusted discharge), and contextual and structural characteristics (rurality, ZIP code–level poverty, ownership, facility type, overall hospital rating, health system membership, teaching hospital status, and COVID-19 Public Health Emergency funding).

For multivariable analyses, predictors were reduced on conceptual relevance and data completeness. Multivariable analysis included: volume/capacity (bed size, inpatient occupancy), patient mix (Medicaid payer mix), financial performance/efficiency (net profit margin, direct patient-care labor cost per adjusted discharge), geography (rurality), and organizational structure (ownership, health system membership, and teaching hospital status)

### Data Collection

Visitor policies were collected through manual review from official hospitals or health-system webpages (October 9–20, 2025); third-party or outdated listings were excluded. If policies were missing or listed only COVID-19–specific restrictions, facilities were contacted by telephone for confirmation. Weekday inpatient visiting start and end times and visitor limits were recorded, prioritizing general medicine/surgery floors when multiple schedules existed. Two authors independently reviewed all policies. Discrepancies were adjudicated by a third author. Hospitals were included if active in both the Centers for Medicare & Medicaid Services dataset fiscal year 2023 and National Academy for State Health Policy fiscal year 2023 datasets, had verifiable visitor policies, and were operational in October 2025. Data from the American Community Survey, United States Department of Agriculture, and the Compendium of U.S. Health Systems were merged using the Centers for Medicare & Medicaid Services Facility ID. Additional variables from other datasets (e.g., poverty, rurality, system membership) were included when available.

### Statistical Analyses

Descriptive statistics were calculated for all variables including distribution of visiting hours (mean, median, SD, range, IQR), and the distribution of visiting hours was visualized using a histogram. The proportion of hospitals within each visiting-hour policy category was also calculated.

Univariate linear regressions estimated associations between each continuous hospital-level predictor and visiting-hour duration; categorical predictors were assessed using one-way ANOVA with Tukey Honestly Significant Difference post hoc tests (27,28). Consistent with a descriptive epidemiologic approach, analyses included univariate associations to maintain transparency in estimation and avoid causal or model-based assumptions that could obscure population-level patterns (29).

A multivariable linear mixed-effects model with log-transformed visiting hours (log[1 + hours]) was used to improve model fit and allow interpretation of coefficients as proportional change. A random intercept for state was included to account for clustering of hospitals within states. Fixed effects included standardized continuous predictors and categorical indicators for ownership, rurality, system membership, and teaching status.

Primary models used standardized (z-scored) predictors to facilitate comparison of effect sizes across variables, whereas secondary models used rescaled predictors for interpretability in natural units (e.g., per $1,000 or per 100 beds). P-values were adjusted using Bonferroni and False Discovery Rate methods.

Nonlinearity was assessed by including linear and quadratic terms for Medicaid payer mix. An ordered logistic mixed-effects model evaluated associations across visiting-hour categories. Odds ratios and confidence intervals were estimated, and p-values were adjusted for multiple testing. A one-way ANOVA with state as the grouping variable was used to assess variance in visiting-hour duration into between-state and within-state components. All analyses were conducted in R 4.5.1. Analyses were restricted to complete cases for all variables included in each model.

### Ethical Considerations

All data is from publicly available government and institutional sources. Only aggregated, non-identifiable information was used. The study was exempt from institutional review. Analyses followed standard principles of transparency, accuracy, and public accountability.

## RESULTS

Four states were selected to capture variation in Medicaid expansion status while keeping comparable population size and hospital capacity. Massachusetts represented a full-expansion state, Wisconsin as a state that expanded Medicaid coverage without the federal match, and Tennessee and South Carolina as non-expansion states (Table 1). Of 350 Centers for Medicare & Medicaid Services hospitals and 333 National Academy for State Health Policy hospitals, the final sample included 318 active hospitals after merging and excluding unmatched, duplicate, or closed facilities (Tables 2-3; Table S1). Several financial and operational characteristics were associated with visiting-hour restrictiveness (Table 4), although models explained limited variance (max adjusted R² = 0.06 for rurality). Mean visiting duration was 14.1 hours/day (SD = 5.07; median = 12; range 0–24; IQR 12–15). Most hospitals had moderately restrictive policies (55.7%), followed by generous (24.8%) and open policies (18.5%); only 1.3% were highly restrictive (Figure 1). Visiting hours clustered between 12–15 hours with a right-bound concentration at 24 hours.

**Figure 1.**
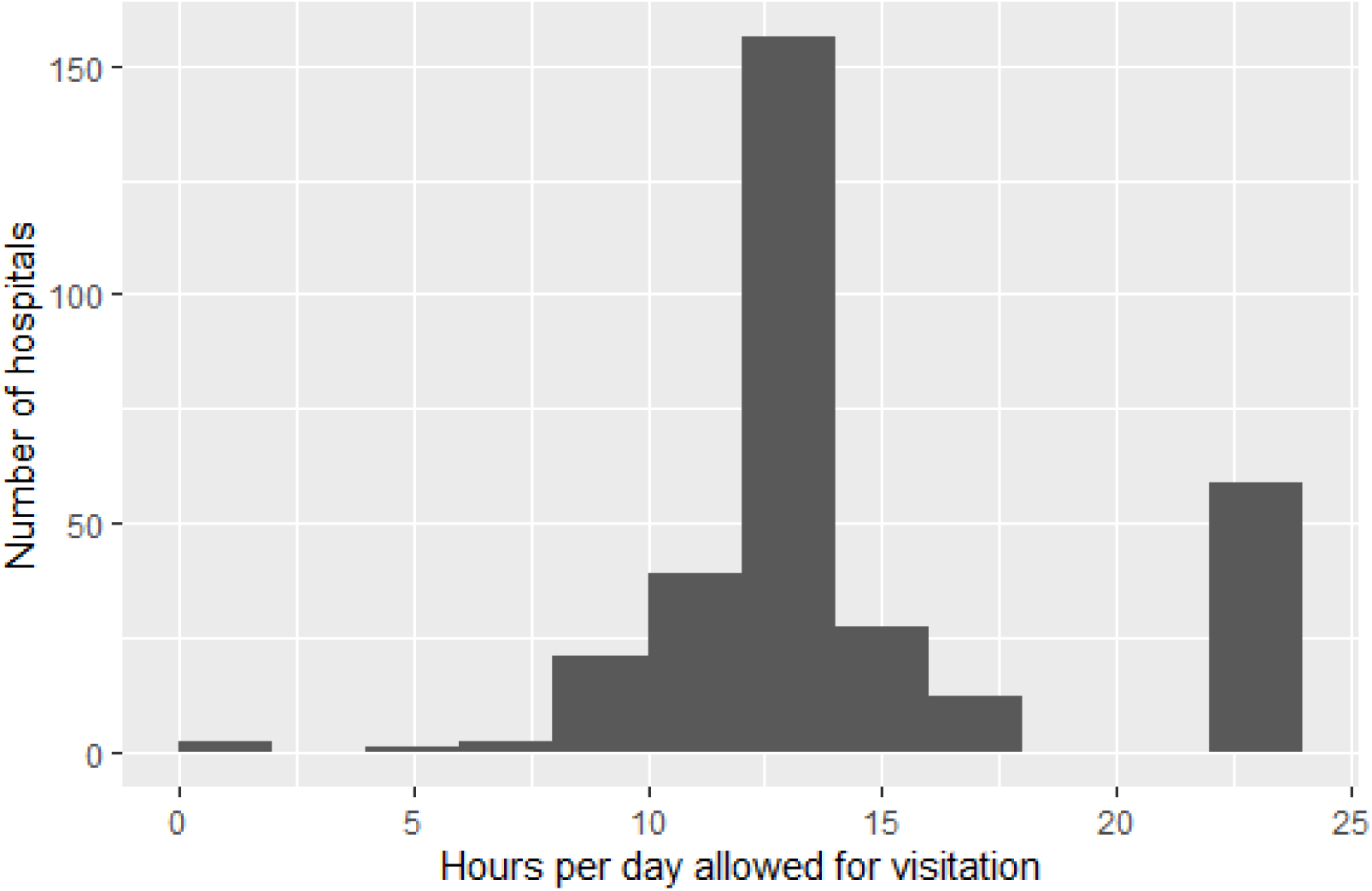
Distribution of Hospital Visiting Hours

**Table 2.**
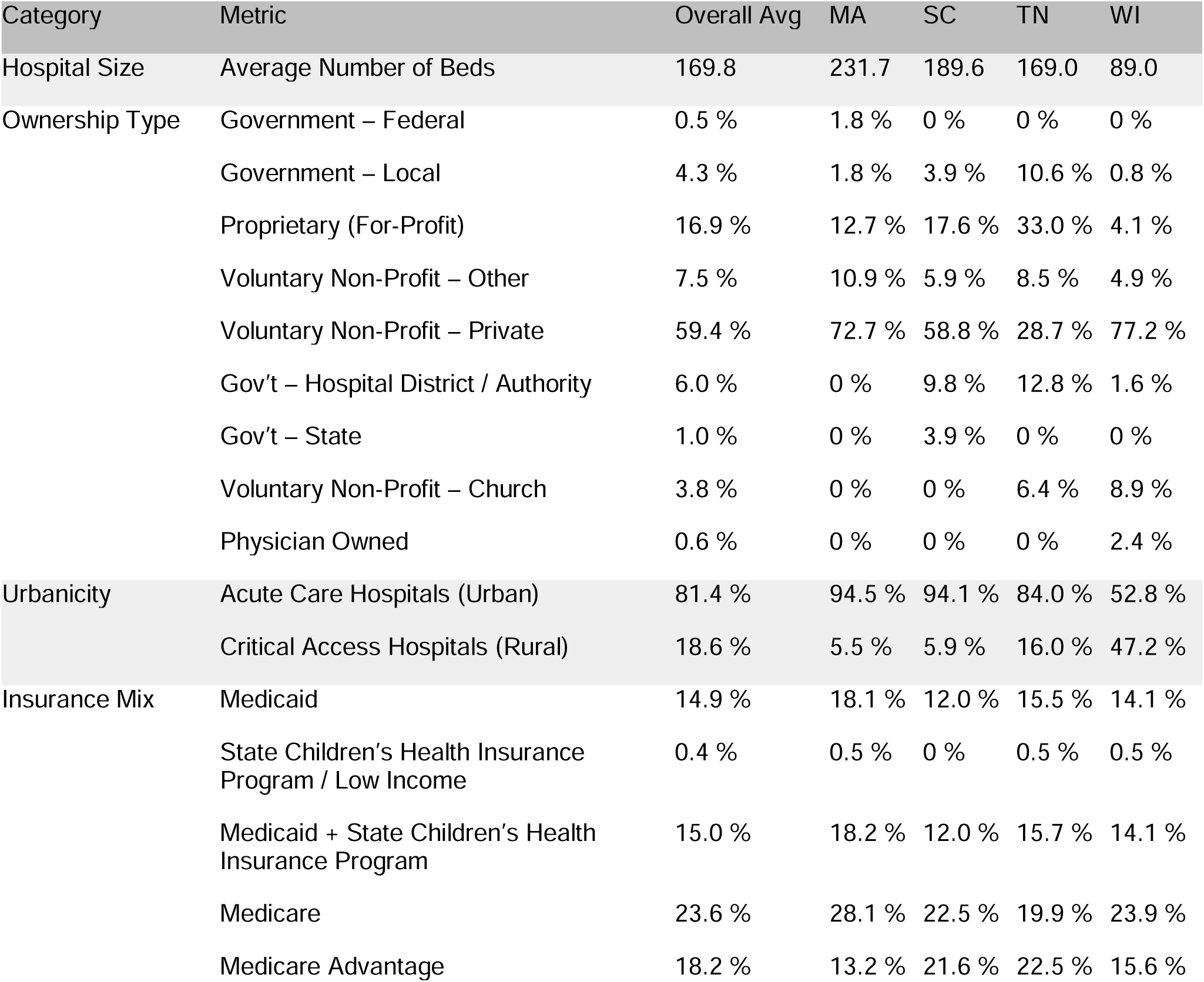
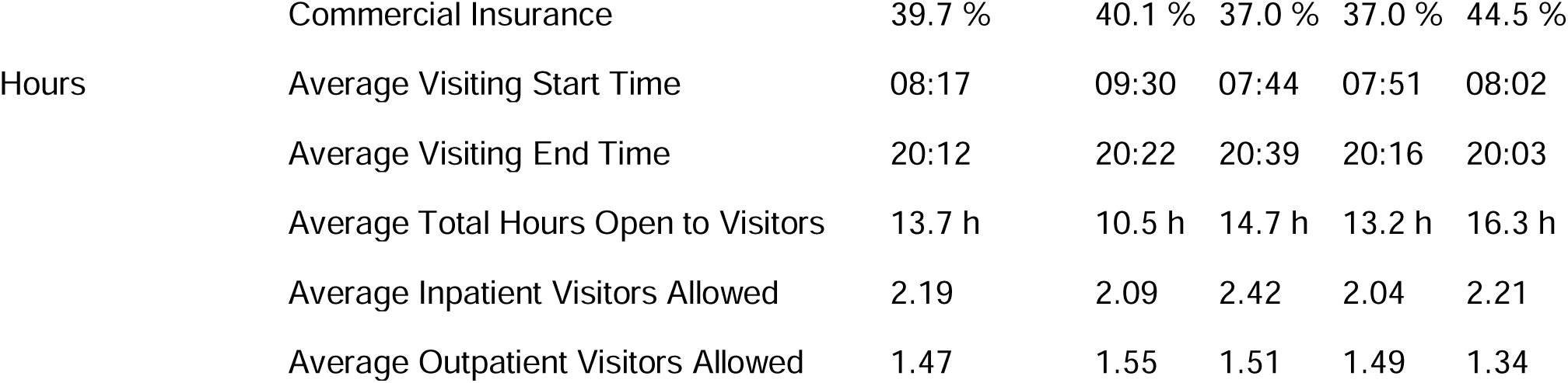
Structural, Ownership, Urbanicity, and Payer-Mix Characteristics of Hospitals by State.

**Table 3.**
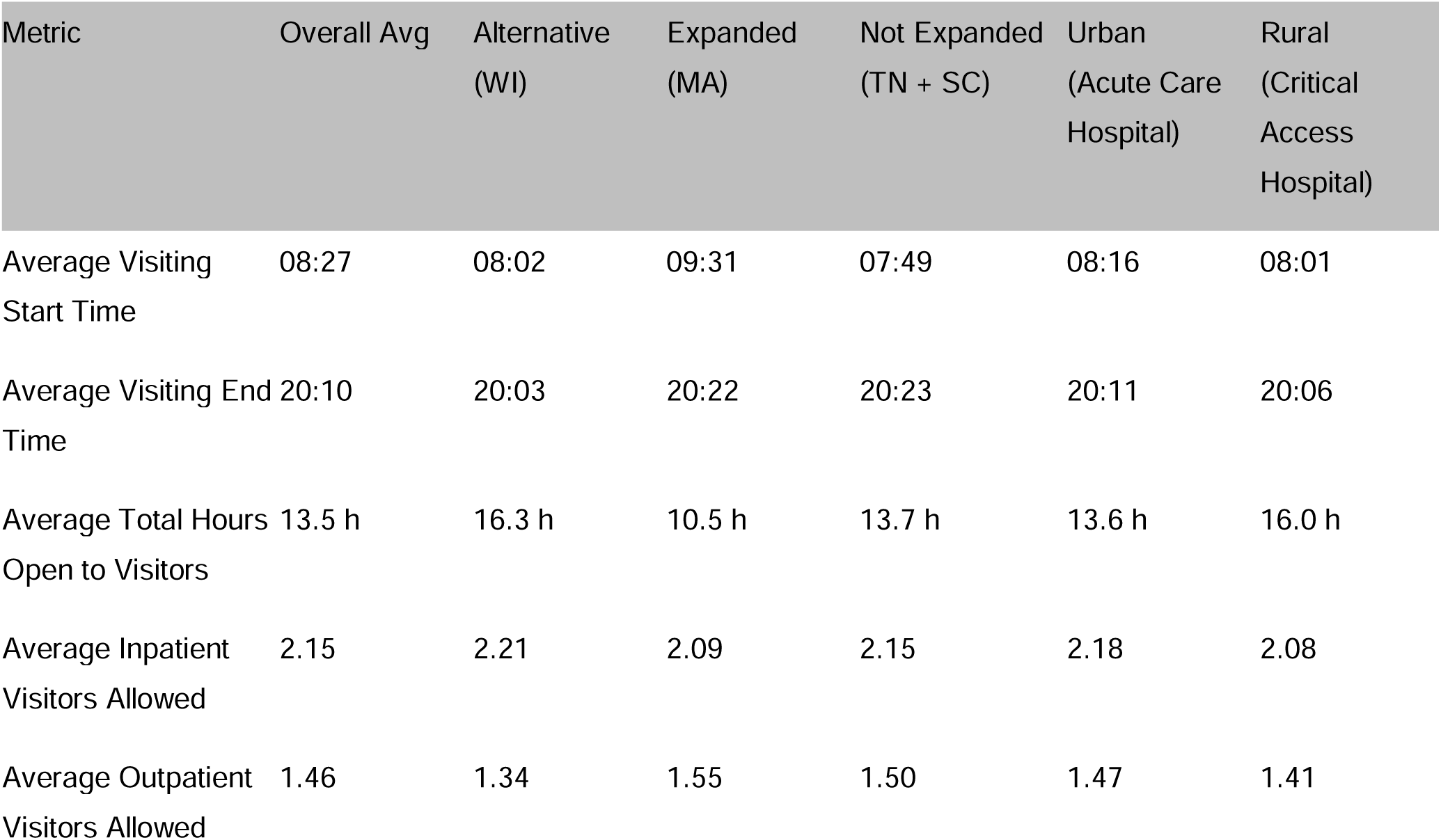
Comparison of Visiting-Hour Policies Between Medicaid Expansion, Non-Expansion, Alternative-Expansion Policy Environments, Urban Acute Care and Rural Critical Access Hospitals.

**Table 4.** Univariate Linear Regression Models Predicting Daily Visiting-Hour Duration from Hospital Financial, Structural, and Labor Characteristics.

| Variable | Estimate ($\beta$) | Std. Error | p-value | Adj. R<sup>2</sup> | N | Min | Max |
| --- | --- | --- | --- | --- | --- | --- | --- |
| Medicaid Payer Mix | −10.843 | 4.628 | 0.020 | 0.014 | 313 | 0.01 | 0.47 |
| Medicaid Operating Profit Margin | −1.583 | 0.407 | <0.001 | 0.044 | 308 | −4.63 | 1.00 |
| Net Profit Margin | −2.222 | 1.050 | 0.035 | 0.011 | 315 | −3.31 | 0.68 |
| Inpatient Occupancy Rate | −4.283 | 1.177 | <0.001 | 0.037 | 317 | 0.00 | 1.03 |
| Bed Size | −0.004 | 0.001 | 0.005 | 0.021 | 318 | 2 | 1,322 |
| Operating Profit Margin | −2.160 | 1.077 | 0.046 | 0.009 | 315 | −2.52 | 0.88 |
| Net Patient Revenue | 0.000 | 0.000 | 0.082 | 0.006 | 315 | \$3,778,174 | \$5,441,708,000 |
| Operating Expenses | 0.000 | 0.000 | 0.064 | 0.008 | 318 | \$7,734,051 | \$6,305,975,000 |
| Net Income (Loss) | 0.000 | 0.000 | 0.235 | 0.001 | 315 | −\$265,931,511 | \$354,430,100 |
| Fund Balance | 0.000 | 0.000 | 0.100 | 0.005 | 318 | −\$646,290,143 | \$5,452,767,000 |
| Hospital Operating Costs | 0.000 | 0.000 | 0.050 | 0.009 | 318 | \$6,010,636 | \$3,087,733,000 |
| Cost-to-Charge Ratio | +3.986 | 1.893 | 0.036 | 0.011 | 318 | 0.07 | 0.96 |
| Net Charity Care Cost (% Expenses) | +2.657 | 7.567 | 0.726 | −0.003 | 304 | −0.19 | 0.43 |
| Uninsured & Bad Debt Cost (% of Expenses) | −6.340 | 18.263 | 0.729 | −0.003 | 316 | 0.00 | 0.12 |
| Medicare Payer Mix | −1.959 | 3.986 | 0.623 | −0.002 | 318 | 0.03 | 0.44 |
| Commercial Payer Mix | +10.202 | 2.536 | <0.001 | 0.046 | 318 | 0.09 | 0.83 |
| Charity Care Payer Mix | −9.642 | 9.964 | 0.334 | 0.000 | 304 | 0.00 | 0.21 |
| Uninsured & Bad Debt Payer Mix | −12.885 | 16.360 | 0.432 | −0.001 | 316 | 0.00 | 0.14 |
| Direct Patient-Care Labor Cost per Full-Time Equivalent | −0.000039 | <0.001 | 0.013 | 0.022 | 238 | \$63,644 | \$181,036 |
| Hospital Operating Labor Cost per Full-Time Equivalent | −0.000042 | <0.001 | 0.022 | 0.018 | 238 | \$61,734 | \$156,946 |
| Adjusted Patient Discharges | −0.000048 | <0.001 | <0.001 | 0.037 | 303 | 169 | 121,146 |
| COVID-19 Public Health Emergency Funding | −0.0000001 | <0.001 | 0.041 | 0.035 | 91 | −\$865,028 | \$60,252,190 |
| Estimated Poverty Level | −0.059 | 0.036 | 0.097 | 0.006 | 305 | 2.3 | 74.0 |
| Rural–Urban<br>Commuting Area Rating | +0.392 | 0.082 | <0.001 | 0.064 | 318 | 1 | 10 |

\* Estimates were rounded using a consistent, magnitude-based precision rule. Coefficients $\geq 0.01$ were rounded to three significant figures. Coefficients $< 0.01$ were rounded to six decimal places, and values that remained zero after six decimal places were rounded to three significant figures. This magnitude-based approach preserves precision across variables measured on different scales.

Higher Medicaid payer mix predicted shorter visiting hours: each percentage increase corresponded to ∼7 fewer minutes per day (β = –10.84, p = 0.02). Higher Medicaid margins also reduced hours, with each point predicting a 1.6-hour decrease (β = –1.58, p < 0.001). Profitability showed the same pattern: higher net profit (p = 0.04) and operating margin (p = 0.046) predicted a 2.2-hour decrease. Higher cost-to-charge ratios, an indicator of higher hospital costs (β = +3.99, p = 0.04), and commercial payer mix (β = +10.20, p < 0.001) were associated with longer visiting hours. Other financial measures were not significant (Table 5).

**Table 5.**
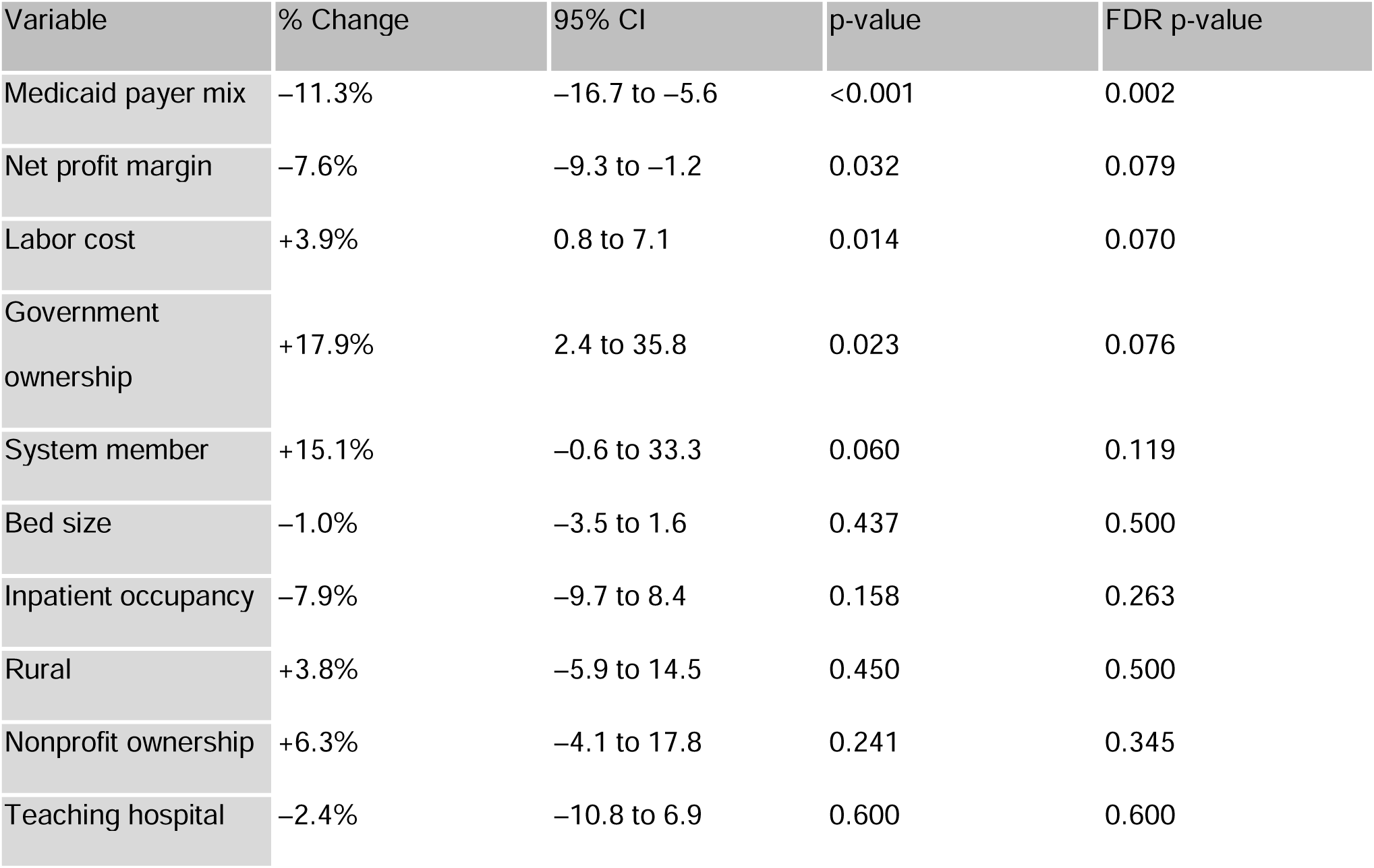
Multivariable Mixed-Effects Model Predicting Daily Visiting-Hour Duration from Hospital Financial, Structural, and Organizational Characteristics.

Capacity and utilization predicted shorter hours. Higher inpatient occupancy (β = –4.28, p < 0.001) and larger hospitals allowed fewer hours: every 100 beds corresponded to 24 fewer minutes (β = –0.004, p = 0.01).

Labor costs showed similar associations. Each $1,000 increase in direct patient-care labor cost per fulltime equivalent predicted ∼2.3 fewer minutes (β = –3.91 × 10⁻, p = 0.013), with total labor cost per fulltime equivalent showing comparable results (β = –4.16 × 10⁻, p = 0.02). Across the observed range ($61,734–$156,946), this equals ∼4 hours difference.

Rurality strongly increased visiting hours: each increasing Rural–Urban Commuting Area level predicted ∼24 more minutes (β = +0.39, p < 0.001), or ∼3.5 hours difference between the most urban and most rural hospitals. Critical Access Hospitals similarly allowed 2.4 more hours (95% CI –3.67 to –1.11; p < 0.001). Zip-code poverty was not significant.

Visiting hours varied by state. Compared with Massachusetts, South Carolina (+4.2 hours; 95% CI: 1.79–6.51, p < 0.001), Tennessee (+2.7 hours; 95% CI: 0.60–4.74, p = 0.005), and Wisconsin (+5.8 hours, 95% CI: 3.77–7.73, p < 0.001) allowed more access. Wisconsin hospitals allowed 3.1 more hours than Tennessee (p < 0.001) but did not differ significantly from South Carolina. State explained 16.4% of variance across hospitals, with 83.6% within-state variation.

In multivariable mixed-effects models (n = 227), Medicaid payer mix was the only predictor consistently associated with visiting-hour restrictiveness after multiple-testing correction (Table 5). Each 10-percentage-point increase was associated with an ∼11.3% decrease in visiting hours (∼1.5 fewer hours/day; p = 0.002). Other associations, including labor cost and government ownership, were not robust after correction. No significant adjusted associations were observed for bed size, occupancy, rurality, nonprofit ownership, or teaching status. A quadratic model indicated a strengthening negative association at higher Medicaid levels (β = –0.0275, p = 0.002). In ordered logistic models, higher Medicaid payer mix (OR = 0.67, p = 0.0256) and net profit margin (OR = 0.63, p = 0.0381) were associated with lower odds of more permissive policies, though these findings were not significant after multiple-testing correction.

## DISCUSSION

This paper evaluated predictors of hospital visitation policy among inpatient general medicine wards within hospitals across four states. In univariate analyses, hospitals with higher Medicaid payer mix and stronger financial margins had shorter visiting hours, whereas greater commercial payer mix, rural location, and smaller size were associated with longer visitation. State-level differences were modest, explaining 16.4% of variation, with most variation occurring within states. In multivariable mixed-effects models, Medicaid payer mix was the only predictor consistently associated with visiting-hour restrictiveness and the only variable that remained statistically significant after multiple-testing correction. Specifically, each 10% increase in Medicaid payer mix was associated with approximately 1.5 fewer hours of visitation per day, with a stronger relationship at higher levels of Medicaid payer mix. Overall, once these factors were considered jointly, most associations attenuated, leaving Medicaid payer mix as the primary driver of visiting-hour restrictiveness.

Although these associations did not persist after adjustment, patterns observed in univariate analyses may still reflect underlying operational and contextual differences (29). Rural hospitals may offer broader hours to accommodate longer travel times, closer community relationships, or local expectations around access. That may be due to lower patient volume, greater operational flexibility, and the ability to transfer complex cases elsewhere. Larger hospitals with higher inpatient occupancy rates, bed count, and more discharges had shorter predicted total visiting hours compared to smaller hospitals. They may adopt more restrictive policies due to the need for more active visitor management because of higher volume of patients (9). Hospitals serving larger Medicaid populations may face staffing or operational constraints that limit flexibility, particularly at high capacity, and their leadership may also have different perceptions about the potential for visitor-related disruptions (30). Prior work suggests that higher Medicaid coverage is associated with decreased patient satisfaction (22,31,32), while hospitals serving more commercially insurance populations may face pressure to offer amenities such as broader visitation hours (12).

Although some financial indicators were associated with visiting hours in unadjusted or partially adjusted models, these relationships were not robust after multiple-testing correction and should be interpreted cautiously (29). Reduced visiting hours correlating with an improved profit margin both from Medicaid and overall may reflect a focus on efficient care to the detriment of non-clinical amenities, if visitation is viewed as an amenity. Hospitals serving larger publicly insured populations might limit non-reimbursable activities such as extended visitation to maintain efficiency, though there is an absence of data showing that extended visitation hours imposes significant costs on hospitals (33,34).

Research shows that improved margins under expansion does not necessarily translate to improvement in care quality in safety-net hospitals (23). Although Medicaid expansion has improved hospital finances by reducing uncompensated care (from 3.9% to 2.3%) within two years of ACA implementation, Massachusetts hospitals still maintained some of the shortest visiting hours in our sample (35,36). Our findings do not suggest that Medicaid expansion worsens visitation policies; rather, expansion may be associated with structural conditions that coincide with more restrictive visitation patterns. Expansion has improved coverage and reduced mortality (35).

While non-expansion states included more rural hospitals, which correlated with broader visitation, most variation occurred within states, disproving our hypothesis that Medicaid expansion status would associate with increased visitation hours. Our findings suggest that state-level factors shape visitation policies less than hospital-level factors such as resource constraints, community relationships, and regional standards. Institutional inertia may also contribute, as visitation policies may persist in the absence of periodic reassessment. Hospitals may independently set visiting-hour policies, largely independent of state influence. Limited state regulations surrounding visitation policies reinforce this theory.

Despite evidence that flexible visitation improves communication and satisfaction, reduces anxiety and delirium, and does not meaningfully increase security incidents or compromise infection control (1,4,5,10,37,38), visiting windows varied widely in our sample. The heterogeneity suggests hospitals balance competing priorities, including patient experience, impact on staff, and operational efficiency, rather than converging on a single optimal policy. One survey of 138 U.S. hospitals visitor policies showed no increase in security calls and the same or fewer visitor-related security issues after switching to open visitation policies (9). This paper also reported that visitor management programs cost on average $44,196 a year, arguing against visitor management representing a financial burden.

Visitor policies have a complex relationship to patients themselves. The available literature shows increased visitor and patient satisfaction with increased visitation policies, largely in the ICU context (39) though with case reports of a hospital system and a large tertiary care hospital providing similar results (10,11). Staff have had concerns about the impact of visitors on the provision of patient care for decades (8), but available meta-analyses working with the limited data available did not find liberalized visitation correlated with worsened clinical outcomes or staff dissatisfaction (40), and the largest RCT did not show increased staff burnout (25). A survey of hospitals focused on visitor management do find concerns of increasing violence within hospital contexts from visitors (9), and an increased Medicaid payer mix may correlate with an increased perceived risk or actual incidence of disruptive behavior from visitors. That same study found no reported increase in security calls with open visitation, and hospitals that reported pre–post data showed the same or fewer visitor-related security issues after switching to open visitation policies (9). Given our findings, further research might focus on hospitals with high Medicaid payer mixes and use qualitative methods to explore why institutions arrived at restrictive or open visitor policies.

## LIMITATIONS

This study had several limitations. First, the study examined only four states, and the nonrandom sampling strategy may restrict generalizability, though we did review all hospitalists with available data within each state. Second, the number of hospitals included was small, reducing the precision of the effect estimates. Third, most financial and administrative data was from fiscal year 2023, whereas visiting-hour policies were collected in 2025, which may introduce temporal misalignment between our data. We believe that relatively few hospitals will have changed their policy during this period. Fourth, the study relied on publicly available administrative data, and as a cross-sectional observational study, analysis should be interpreted as associative rather than causal.

## CONCLUSION

Reviewing visitation policies from acute care hospitals across 4 states, this study found a complex relationship between policies and hospital location, payer mix, size, and financial profile, with the Medicaid payer mix being the sole statistically significant variable on multivariate analysis. The range of visitor hours found suggests potential opportunities to expand patient-centered approaches to visitation while monitoring for impacts on hospital finances, staff, and clinical care.

## Data Availability

The datasets analyzed during the current study are publicly available from the following sources: (1) the Centers for Medicare & Medicaid Services Hospital General Information dataset (available at: https://data.cms.gov/provider-data/dataset/xubh-q36u); (2) the National Academy for State Health Policy Hospital Cost Tool (available at: https://www.nashp.org/hospital-cost-tool/); (3) the U.S. Census Bureau American Community Survey 2023 Table S1701 “Poverty Status in the Past 12 Months” (available at: https://www.census.gov/) (4) the U.S. Department of Agriculture Rural-Urban Commuting Area Codes (available at: https://www.ers.usda.gov/data-products/rural-urban-commuting-area-codes/); and (5) the Agency for Healthcare Research and Quality Compendium of U.S. Health Systems (available at: https://www.ahrq.gov/chsp/data-resources/compendium-2023.html). The manually compiled dataset of hospital visiting hours used in this analysis, including the code linking these sources, is available from the corresponding author on reasonable request. Visitor policy data collection was conducted between October 9 and October 19, 2025

## ACKNOWLEDGEMENTS

FUNDERS

The authors did not receive any internal or external funding.

## PRIOR PRESENTATIONS

This data has not been presented in paper, abstract, or poster form in any other settings.

## CLINICAL TRIAL NUMBER

Not applicable.

## AUTHORSHIP CONTRIBUTION

All authors agree to be accountable for all aspects of the work.

Conception and study design: JCH, LTAB

Data acquisition: CYG, CC, LC

Data verification: CYG, CC, JCH

Statistical analysis: CYG, LTAB

Manuscript preparation: CYG

Manuscript editing and review: CYG, JCH, LTAB

## LIST OF ABBREVIATIONS

ACA: Affordable Care Act
ANOVA: Analysis of Variance
MA: Massachusetts
SC: South Carolina
TN: Tennessee
WI: Wisconsin

## ETHICS APPROVAL AND CONSENT TO PARTICIPATE

This research used solely publicly available data and did not use any patient level data; as such, it was exempt from institutional review.

## SUPPLEMENTARY MATERIALS

**Table S1.** Hospitals Excluded from the Analytic Sample, by State and Reason for Exclusion.

| State | Hospitals Excluded | Excluded Due to Closure | Excluded Due to Unmatched National Academy for State Health Policy Record | Average Opening Time | Average Closing Time |
| --- | --- | --- | --- | --- | --- |
| Massachusetts | 10 | 4 | 6 | 08:00 | 21:12 |
| Tennessee | 6 | 0 | 6 | 05:20 | 22:00 |
| South Carolina | 6 | 0 | 6 | 06:00 | 17:00 |
| Wisconsin | 5 | 0 | 5 | 05:00 | 20:16 |
| Total | 27 | 4 | 23 | ≈ 06:30 | ≈ 21:09 |

**Table S2.** Categories of Hospitals Excluded from Analysis.

| State | Veterans Affairs Hospitals* | Unmatched Acute Care Hospitals | Children's Hospitals* | Unmatched Critical Access Hospitals | Total Excluded |
| --- | --- | --- | --- | --- | --- |
| Massachusetts | 3 | 3 | 3 | 1 | 10 |
| Tennessee | 4 | 0 | 2 | 0 | 6 |
| South Carolina | 2 | 3 | 1 | 0 | 6 |
| Wisconsin | 3 | 0 | 2 | 0 | 5 |
| Total | 12 | 6 | 8 | 1 | 27 |

\* All Veterans Affairs and children's hospitals were excluded a priori due to non-comparability with the Centers for Medicare & Medicaid Services dataset.

**Table S3.**
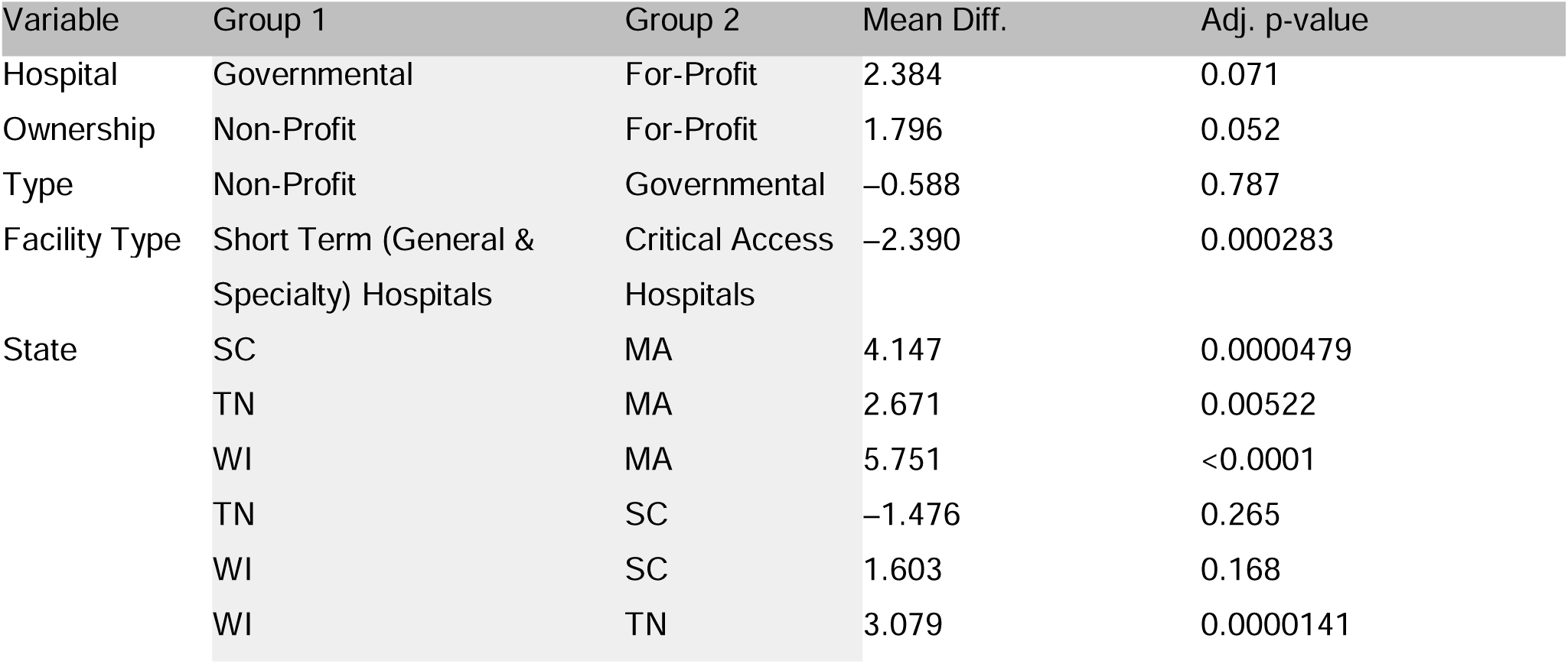
Variance and Tukey Post Hoc Comparisons Across Hospital Ownership, Facility Type, and State.

**Table S4.** Pairwise Comparisons of Visiting-Hour Duration Across Centers for Medicare & Medicaid Services Overall Hospital Quality Ratings.

| Rating Category (Comparison A) | Rating Category (Comparison B) | Mean Difference (hours) | Adj. p-value |
| --- | --- | --- | --- |
| 2 | 1 | 1.702 | 0.959 |
| 3 | 1 | 2.496 | 0.802 |
| 4 | 1 | 1.593 | 0.966 |
| 5 | 1 | 2.648 | 0.811 |
| Not Available | 1 | 4.347 | 0.225 |
| 3 | 2 | 0.794 | 0.958 |
| 4 | 2 | −0.109 | 1.000 |
| 5 | 2 | 0.946 | 0.971 |
| Not Available | 2 | 2.645 | 0.0367 |
| 4 | 3 | −0.903 | 0.888 |
| 5 | 3 | 0.152 | 1.000 |
| Not Available | 3 | 1.851 | 0.156 |
| 5 | 4 | 1.055 | 0.941 |
| Not Available | 4 | 2.754 | 0.00641 |
| Not Available | 5 | 1.699 | 0.633 |

